# High throughput SARS-CoV-2 variant analysis using molecular barcodes coupled with Next Generation Sequencing

**DOI:** 10.1101/2021.05.27.21257726

**Authors:** Lyora A. Cohen-Aharonov, Annie Rebibo-Sabbah, Adar Yaacov, Roy Z. Granit, Merav Strauss, Raul Colodner, Ori Cheshin, Shai Rosenberg, Ronen Eavri

**Affiliations:** Barcode Diagnostics Ltd., Nazareth, Israel; Laboratory for Computational Biology of Cancer, Sharett Institute for Oncology, Hadassah - Hebrew University Medical Center; The Wohl Institute for Translational Medicine, Hadassah – Hebrew University Medical Center; Microbiology Laboratory, Emek Medical Center, Afula, Israel; Internal Medicine E, Emek Medical Center, Afula, Israel

**Keywords:** Next-generation sequencing, COVID-19, molecular barcodes, SARS-Cov-2 diagnosis, high-throughput detection, variant analysis

## Abstract

The identification of SARS-CoV-2 variants across the globe and their implications on the outspread of the pandemic, infection potential and resistance to vaccination, requires modification of the current diagnostic methods to map out viral mutations rapidly and reliably. Here, we demonstrate that integrating DNA barcoding technology, sample pooling and Next Generation Sequencing (NGS) provide an applicable solution for large-population viral screening combined with specific variant analysis.

Our solution allows high throughput testing by barcoding each sample, followed by pooling of test samples using a multi-step procedure. First, patient-specific barcodes are added to the primers used in a one-step RT-PCR reaction, amplifying three different viral genes and one human housekeeping gene (as internal control). Then, samples are pooled, purified and finally, the generated sequences are read using an Illumina NGS system to identify the positive samples with a sensitivity of 82.5% and a specificity of 97.3%. Using this solution, we were able to identify six known and one unknown SARS-CoV-2 variants in a screen of 960 samples out of which 258 (27%) were positive for the virus.

Thus, our diagnostic solution integrates the benefits of large population and epidemiological screening together with sensitive and specific identification of positive samples including variant analysis at a single nucleotide resolution.

## Introduction

The severe acute respiratory syndrome coronavirus 2 (SARS-CoV-2) is an RNA virus that causes the coronavirus disease 2019 (COVID-19). The virus was first identified in Wuhan (China) in December 2019, spread rapidly and was declared a pandemic in March 2020 by the World Health Organization. Today, more than a year later, COVID 19 has already resulted in over 160 million confirmed infection cases and over 3 million deaths across the world.

COVID-19 symptoms are varied, ranging from none to severe. Many patients develop moderate pneumonia and in the most critical cases respiratory failure and multiorgan disfunction occur. The world average mortality is 2.2%.

Prevention of viral transmission is essential as COVID-19 spreads through the respiratory route. Transmission prevention is achieved through social distancing, masks and hand washing in addition to early identification of potential infected individuals by different diagnostic methods. SARS-CoV-2, a positive-sense single-stranded RNA virus is a highly pathogenic member of the coronavirus family ^1^. The sequence of its whole genome was published (GenBank no. MN908947), encoding 9860 amino acids. It is composed of genes that express both structural and nonstructural proteins.

S-, E-, N- and M- genes encode for structural proteins (spike, envelope, nucleocapsid and membrane proteins respectively), ORF region encodes for non-structural proteins as RNA-dependent RNA polymerase (RdRp) or papain-like protease ^2^.

Several methods have been developed in order to assess the infection status of individuals. Most of them rely on the detection of viral RNA.

The gold-standard method for diagnosis of COVID-19 is real-time reverse transcription polymerase chain reaction (one step rRT-PCR) ^3^, which detects the presence of viral RNA fragments. The test is performed on nasopharyngeal swabs or saliva samples ^4^ and results are generally delivered within 24-48 hours. A number of companies have developed real time PCR diagnostic tests for SARS-CoV-2 detection. These methods vary in sensitivity and in the ability to detect different parts of the viral genome.

However, real time PCR is not a high throughput method, the number of amplicons that can be detected are limited, and fluorescent probes and special reagents are needed in order to increase the sensitivity of the method. Moreover, as the global spread of the virus continues despite the use of preventative measures and positive effects of vaccination^5^, SARS-CoV-2 mutations generate different viral variants, some with the potential to become more virulent and contagious or resistant to vaccination. It is therefore, essential to develop accurate and cost-effective diagnostic methods which can provide mass screening abilities combined with sequence-specific mutation analysis.

Next-generation diagnostic solutions, which are based on the current PCR test coupled with next generation sequencing (NGS) offer major advantages, such as significantly larger scale testing (several hundred-fold scale up versus the currently available test), reduced cost per test, reduced amount of reagents per test and potential readout of viral sequence variation.

In NGS-based methods, individual samples can be uniquely labeled with molecular barcodes and multiple fragments can be amplified in parallel and pooled together for high throughput detection and processing of thousands of specimens together. A few different protocols have been developed ^6,7^ and one of them has even received an Emergency Use Authorization from the FDA ^8^. As next-generation sequencers are now widely available, including inside hospitals, this platform offers a key solution for COVID-19 mass detection worldwide.

Here we propose a solution that allows high throughput testing by barcoding and pooling test samples, up to hundreds at a time, in one well (as illustrated in Figure 1). The protocol combines multiplexed PCR, patient specific molecular barcoding, pooling, NGS, bioinformatics and machine learning analysis, for identification of positive individuals by detection of three different viral genes and one human gene (as internal control) from a pool of thousands of specimens, enabling sensitive and accurate molecular-based diagnosis of COVID-19. Using this method, we have identified six known and one new viral variants in a mass screening analysis of 960 samples. The test can prove vital in the identification of various viral variants as part of population and epidemiological screening.

**Figure 1:**
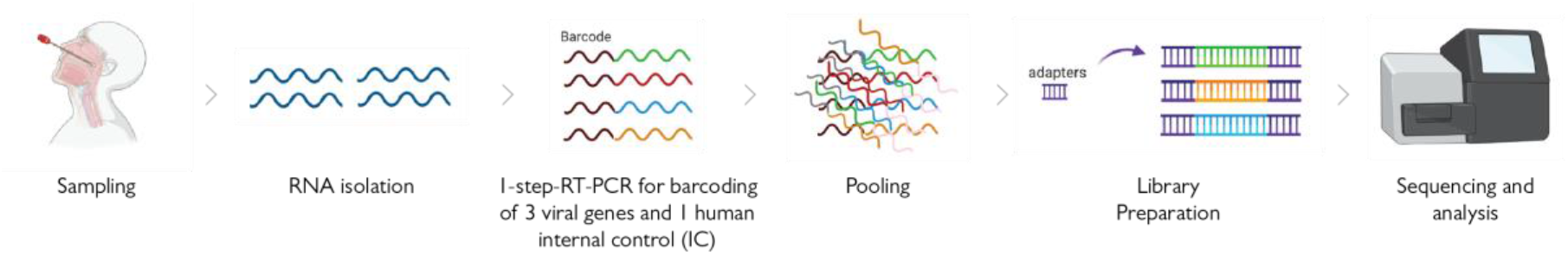
A schematic showing the overall workflow, including viral RNA extraction, one-step reverse transcription-PCR using barcoded-specific primers, and next-generation sequencing to quantify the amounts of barcoded amplicons.

## Results

### Barcodes

A bank of 2133 unique 10-bases and about 21,000 unique 12-bases barcodes was generated. For the purpose of this research, the most distinguishable 96 barcodes were selected for the experiments (see Methods).

### Pilot

Positive and negative SARS-CoV-2 standards (Bio-Rad) were run as a pilot with 17 barcodes and a multiplex of two viral genes (N1 and E) and one human internal control (RNaseP). Following multiplexing, amplicons were pooled. A single library was prepared and ran on a MiSeq (Illumina) instrument.

Following demultiplexing, number of reads for each barcode was obtained. RNase P reads were used for internal control and were required to be positive in order to consider the result as valid. The viral genes were used for the diagnosis and detection of SARS-CoV-2 specifically in each specimen. For each barcode, the associated viral reads were counted and the results are presented in Figure 2a. A clear distinction was observed between positive and negative samples.

**Figure 2:**
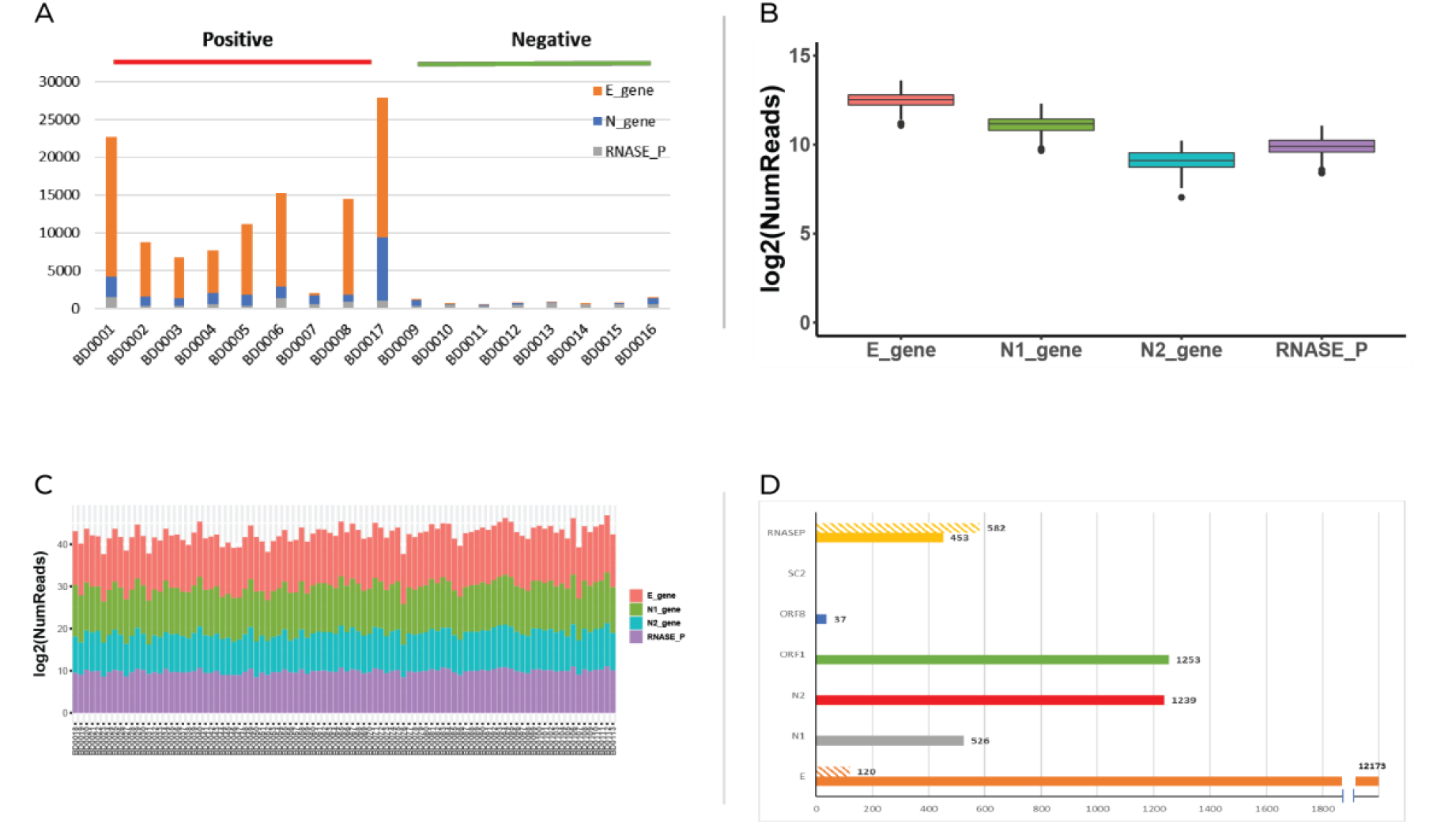
A. 17 clinical RNA samples (9 positive and 8 negative) analyzed by molecular barcoding of 2 viral genes and one internal control human gene and Next-Generation Sequencing. B and C. Stability of each of the 96 barcodes used in the experiment. The same amount of positive standard was used with all the barcodes for comparison. Count of reads is presented in boxplots for each target (E, N1, N2 and RNASE P) (B) and in columns for each barcode (C) D. Analysis of 6 viral genes and one human internal control (plain column- positive standard, striped column- negative standard).

### Barcode stability

Following this experiment, the stability of 96 barcodes was tested using positive SARS-CoV-2 standard as an identical template for each barcode. Furthermore, an additional viral region (N2) was amplified and added to the multiplex. The stability of 96 barcodes is presented in Figure 2b and c. No specific influence of the barcode sequence was noticed. Moreover, the N2 viral fragment added additional information regarding the viral identification and can provide an improvement in diagnosis of the specimens.

### New genesss

In order to improve specificity and sensitivity, additional viral regions for barcoded amplification were examined. Each new set of primers was first tested on positive and negative SARS-CoV-2 standards. The count of reads is presented in Figure 2d. Human RNase P served as the internal control gene. A threshold was set to determine the genes that should be part of the multiplexes. This threshold represents the minimal number of reads of positive samples for a specific target in order to include this specific amplified amplicon in the multiplex assay. As the SC2 assay failed to yield any reads for both positive and negative templates, it was not comprised in the further multiplexes. Additionally, we observed a weaker signal for ORF8 in comparison to ORF1. Thus, N1, N2, E, and ORF1 were selected as the multiplex target fragments for the next step.

### Multiplex optimization

In order to choose the most effective combination of genes targeted, the following multiplexes were tested on 96 clinical RNA specimens for optimization (Figure 3). Multiplex 1 included viral genes N1, N2 and E; multiplex 2 included viral genes N1, N2 and ORF1; and multiplex 3 included viral genes N1, E and ORF1. Human RnaseP was included in all three mixes as internal control.

**Figure 3:**
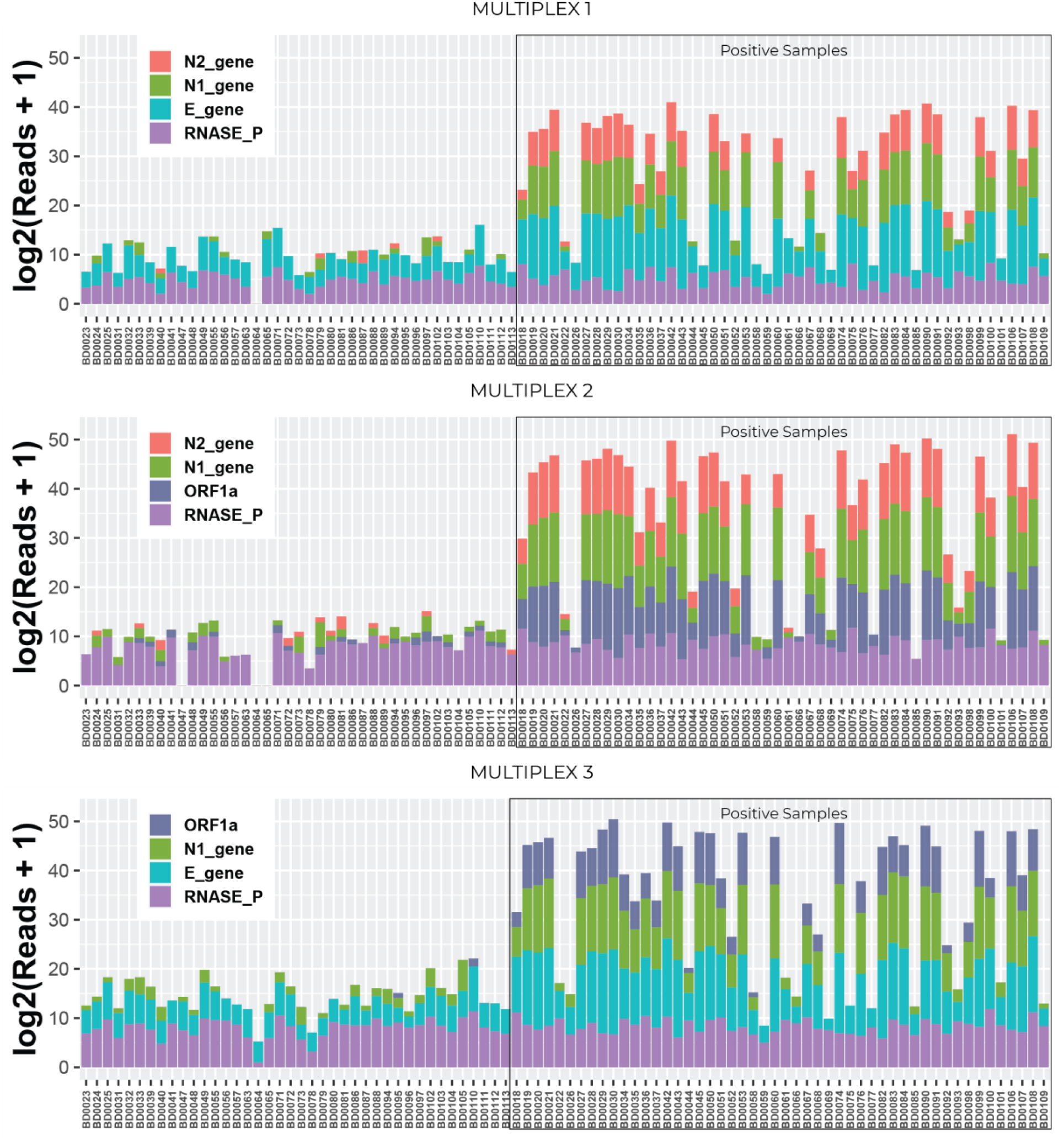
Multiplex optimization. Three different multiplexes including three viral genes among N1, N2, E and ORF1a genes, and including one human internal control gene RNAseP were tested separately with the same clinical specimens in order to compare between them and determine the optimal combination to be use for the test.

Our results demonstrate that when genes N1 and N2 are multiplexed together, a non-specific DNA fragment is generated due to the physical proximity of the two amplicons (unshown data). This 944bp DNA fragment was formed as the elongation product of the N1 forward primer and N2 reverse primer. As this fragment is added to the library preparation and run together with all the amplicons, it can lead to a competition in the NGS run and to a lower number of reads that are obtained from the fragments of - interest.

### Detection of SARS-CoV-2 in a 960 samples pool

In order to scale up our technology, 960 clinical RNA specimens were tested using multiplex 3. Amplicons were pooled (each plate separately), libraries were prepared for each plate with a different index and all ten libraries were sequenced together using the Next-Seq instrument (illumina). Following demultiplexing, obtained sequences were analyzed and positive samples were identified using the unique and specific barcodes (Figure 4a). The number of viral reads in our pool was in negative correlation with the Ct number obtained by regular real-time PCR. When the Ct was high, the number of viral reads in the pool were low (Figure 4b). Our results indicate that at a Ct higher than 30 the number of viral reads is similar between positive and negative specimens.

**Figure 4:**
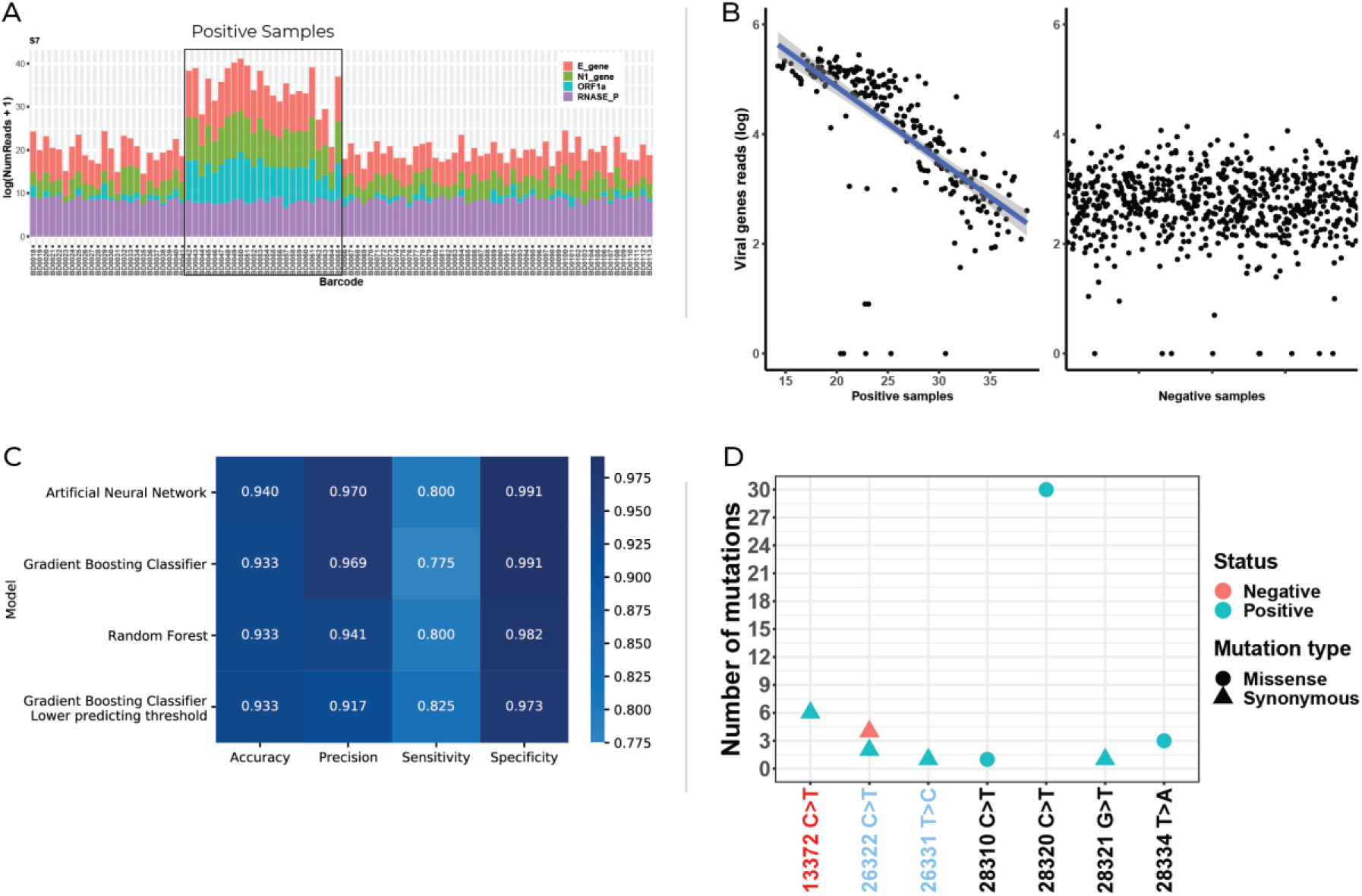
A. Bar charts from one representative plate of total sequencing reads from specimen tested. B. Total viral reads in positive and negative specimens. Positive X axis shows the Ct number obtained from the amplification of a fragment from N gene. C. Heat map showing accuracy, precision, sensitivity and specificity using four different machine learning algorithms. D. A scatter plot showing all detected genomic variants within the 960 samples tested. 6 known SNPs and one heretofore unreported SNP were found.

Using a machine learning classification model, results were obtained with an accuracy of 93.3%, precision of 91.7%, sensitivity of 82.5% and specificity of 97.3% (Figure 3c) in a validation set. We further trained a model defining Ct<30 (N-gene) specimens as positive. The sensitivity was 100% with 98.5% specificity and a positive predictive value (PPV) of 94.7% in a validation set.

### Variant detection

In addition to the detection of the virus with high specificity and sensitivity (especially for Ct <30), SARS-CoV-2 barcoded pooling provides a platform for detection of single nucleotide mutations (Figure 4d) in pre-determined gene regions. We have sequenced 3 viral fragments for detection of the virus in all 960 specimens. Following NGS analysis of the multiplexes, 7 single nucleotide variants were identified. By comparing the observed mutations to SARS_CoV-2 databases using Blastn in betacoronavirus genbank - https://blast.ncbi.nlm.nih.gov/) we have identified six known and one newly found missense mutation. Among the 258 SARS-CoV-2 PCR-positive samples, 30 carried the same N-gene missense mutation and 6 carried an additional synonymous substitution in ORF1a (Figure 4d). Eight PCR-positive and two PCR-negative samples were found with the five remaining single nucleotide variants, one variant per sample.

## Discussion

SARS-CoV-2 (severe acute respiratory syndrome coronavirus 2), a beta coronavirus, is the novel coronavirus that caused the COVID-19 outbreak originating in Wuhan, China in 2019. This virus has become a worldwide pandemic requiring immense and rapid viral detection methods throughout the world. The classic method for detection of infection involves sampling nasopharyngeal swabs from patients in order to detect viral infection. The swab is placed in transfer buffer and then lysis buffer is added to lyse the potentially infected cells. RNA is extracted from the solution then subjected to a one-step RT-PCR reaction. The RNA is first converted to cDNA by reverse transcription which is immediately followed by Multiplex quantitative Real time PCR (qPCR) with primers and a probe specific to the virus to detect viral presence.

Generally, an internal control is added in order to eliminate the possibility of false negative results and includes detection of a human gene as a proxy for host RNA content. However, some assays only amplify an exogenic internal control gene (added during the extraction step) which provides evidence for the validity of the qPCR reaction yet, is not reliable enough to exclude false negatives.

Here we describe a method we developed for combined screening of an infectious agent and simultaneously determine the presence of a genomic variant in a population, by using a DNA-barcoded sample from each subject in a population. The method includes performing one-step reverse transcription and amplification of RNA extracted from the samples with barcoded primers, pooling and sequencing the obtained amplicons, and identifying the infected individual out of all tested samples in the pool via the subject-specific barcode. The different primers target three viral genes as well as one human endogenous gene. The detection of an endogenic human internal control assay that is specific to each specimen, even though they are sequenced as a pool, is of extreme importance and represents one of the major advantages of this technique.

As the virus has spread globally, it has naturally acquired thousands of mutations. Most of these are deleterious or neutral, however, some confer selective advantage and have given rise to many viral variants that became the dominant viral strain in many countries. Some of these variants as they have been shown to increase infection rate, cause higher mortality and most importantly may evade the immune response obtained by vaccination ^17^. It is therefore essential that health care authorities collect evidence on the extent and spread of such viral variants within the population. However, currently there are no cost effective and non-overly labor reliant methods for mass screening of SARS-CoV-2 variants in an entire population.

Our method utilizes amplified cDNA produced from the infectious agent and at least one Single Nucleotide Polymorphism (SNP) in a sequencing read, to determine via the subject-specific barcode whether the subject is infected and simultaneously enables the identification of a genomic variant. The method presented here can potentially be modified using primers designed upstream of the variable S gene to enable the detection of the known variants (UK-B.1.1.7 lineage, South Africa-B.1.351 lineage, Brazil-P.1 lineage, NY-B.1.526 lineage) whose sequences are mutated in this specific locus (Spike gene).

In conclusion, in comparison to the gold standard real time PCR method, that identifies amplified cDNA produced from the infectious agent and is only able to provide a binary answer (SARS-CoV-2 positive or negative), the method presented here, enables specific diagnosis of SARS-CoV-2 variants within a population of thousands. As this solution can either provide a stand-alone identification method for SARS-CoV-2 or be integrated with the current RT-PCR viral test, we propose adopting this technique to large population and epidemiological screening practice.

## Materials and methods

### Clinical specimens

Oro- and nasopharyngeal swabs were collected from patients by trained health workers with personal protective equipment and directly placed into e-swab tubes (COPAN) for conservation and transport.

### RNA extraction

Following sampling and storage of the oro- and nasopharyngeal swabs, nucleic acids were isolated and purified from swab specimens using STARMag 96 × 4 Universal Cartridge Kit (Seegene) on Star or Starlet instruments (Hamilton) following the manufacturer’s instructions.

### Primer sequences

The targets analyzed were amplified using previously reported sets of known primers, while unique barcodes were added to the forward primer to enable pooling of the products before sequencing. These primer sequences are summarized in Table 1.

**Table 1:**
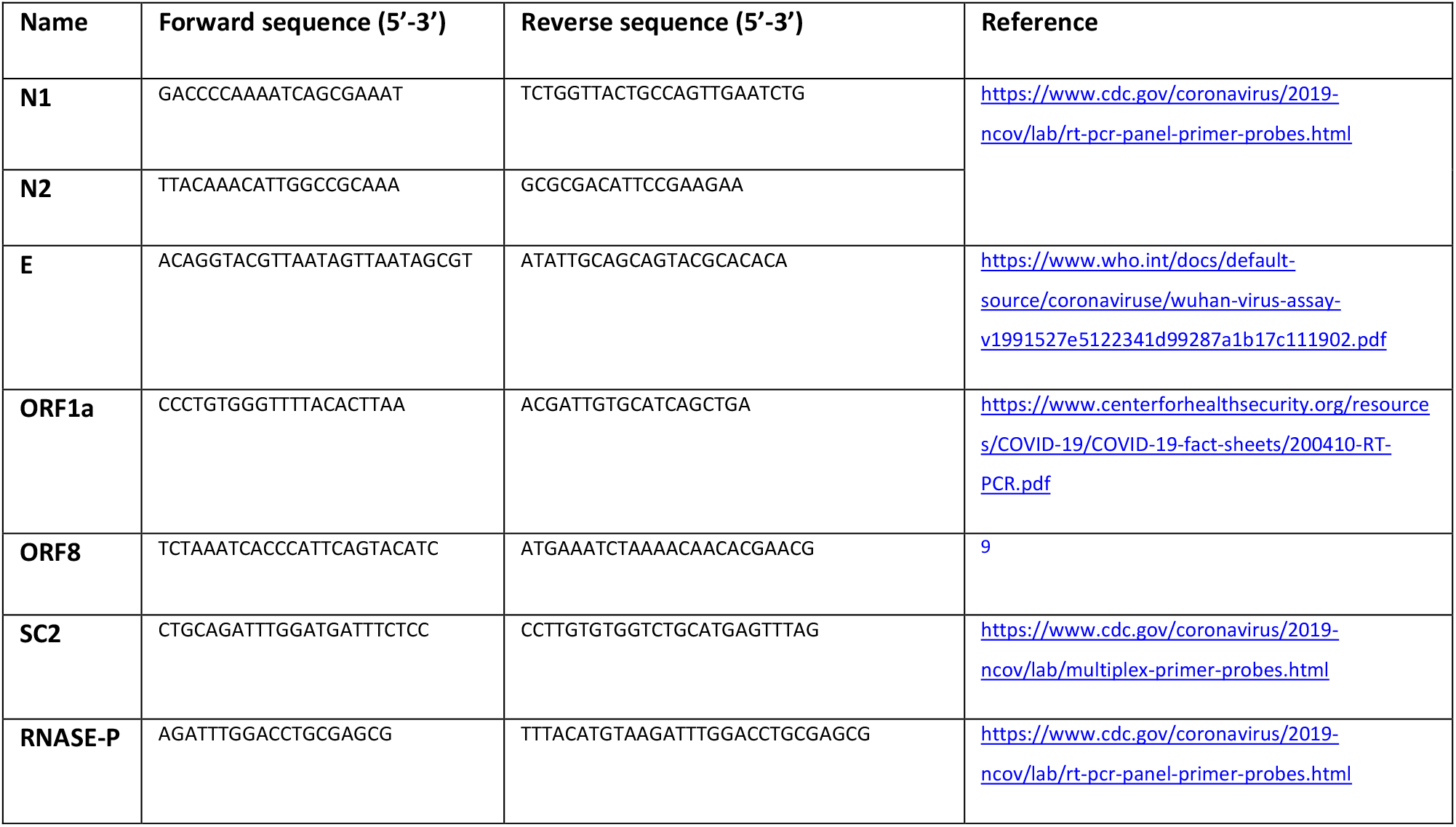
Primers sequences

The complete table of primers including barcodes appears in the supplementary data of this publication.

### Singleplex RT-PCR

The purified nucleic acids were reverse transcribed into cDNA and amplified using Qscript XLT one-step RT-PCR (Quantabio) and specific primers for each viral and human amplicons. Each forward primer included a 10-nucleotide barcode at the 5’-end, which was unique for each sample tested and enabled pooling of the PCR products prior to sequencing. The total volume of reaction mix (25 µl) contained the following: 12.5 µl of 2X concentrated ToughMix; 0.4 µM reverse primer, 0.4 µM barcoded forward primer; 1 µl of 25X qScript XLT and 5 µl of SARS-CoV-2 positive and negative standard (BioRad). The one step RT-PCR was conducted in a C1000 touch thermocycler (BioRad) as per the following: cDNA Synthesis (48°C, 20 min), Initial denaturation (94°C, 3 min), PCR cycling -25 cycles (Denaturation: 94°C, 15s; Annealing: 60°C, 40s; Extension: 72°C, 30s).

### Multiplex RT-PCR

The purified nucleic acids were reverse transcribed into cDNA and amplified using Qscript XLT one-step RT-PCR (Quantabio) and specific primers for 3 viral amplicons (N1, N2, E in multiplex 1; N1, N2 and ORF1 in multiplex 2; and N1, E and ORF1 in multiplex 3) and one human (RNaseP as internal control). Each forward primer included a 10-nucleotide barcode at the 5’-end, which is unique for each individual tested (the barcode generation is described in the next paragraph). The total volume of reaction mix (25 µl) contained the following: 12.5 µl of 2X concentrated ToughMix; 0.4 µM each reverse and barcoded forward primer; 1 µl of 25X qScript XLT and 8 µl of the RNA sample. The one step RT-PCR was conducted in a C1000 touch thermocycler (BioRad) as per the following cDNA Synthesis (48°C, 20 min), Initial denaturation (94°C, 3 min), PCR cycling -25 for pool of 96 specimens /35 cycles for pool of 960 specimens (Denaturation: 94°C, 15s; Annealing: 60°C, 40s; Extension: 72°C, 30s).

### Barcode generation

Sequences of unique ten-base barcodes were created using DNA barcodes R package ^10^. Barcodes were generated in such a manner that they were at least three Sequence-Levenshtein distance apart. In addition, to meet the required chemical properties, sequences that contained triplets that were self-complementary or that showed an unbalanced GC ratio were filtered out. Based on distance metrics between each pair of barcodes, most distinguishable 96 barcodes were selected for the experiments based on the distance between every possible pair among the 96 barcodes.

### Library preparation and next generation sequencing

All the 96 separate reactions of one plate were pooled together. Pooled PCR products were purified using MiniElute Spin Columns (Qiagen) according to manufacturer’s instructions.

Purified PCR amplicons were used as input DNA to generate Illumina-compatible sequencing libraries using NEBNext® Ultra™ II DNA Library Prep Kit for Illumina® (New England BioLabs) according to the manufacturer’s instructions (fragmentation and size-selection were not required). Final sequencing libraries were sequenced on the Illumina Miseq sequencer (Illumina) using 2×150 bp paired-end reads (Micro kit).

A pool of 960 clinical samples was tested using the technology. Ten 96 plates with 96 different specimens (about 25% positive) were amplified using the barcoding-multiplex PCR. Wells from each plate were pooled together and a library was prepared using unique index. All ten libraries were sequenced together using the Next-Seq instrument (illumina).

### Next Generation Sequence analysis

#### Reads processing pipeline

First, a raw FASTQ file was demultiplexed by DNA-barcodes using FASTX barcode splitter, i.e., splitting into numerous files based on DNA-barcode matching, one FASTQ per barcode (FASTX-toolkit version 0.0.14, http://hannonlab.cshl.edu/fastx_toolkit/). One mismatch ^10^ between the known and the observed sequence of a barcode was allowed. Then, each barcode specific FASTQ file was trimmed using FASTX trimmer, leaving 41 bases of each read, from the 11th base to the 51st base, representing a part of the designated primers. The trimmed reads were aligned to N (nucleocapsid phosphoprotein), E (envelope protein) and ORF1ab (ORF1a; ORF1b polyproteins) SARS-CoV-2 genes and human RNASE P (from NCBI Genbank) using BBMap version 38.18, which is a short-reads aligner for DNA or RNA-seq data (http://sourceforge.net/projects/bbmap/). Finally, the set of reads were quantified using Salmon version 0.14.2 ^11^.

#### Variant calling pipeline

For the purpose of variant calling, the barcodes sequences were trimmed at the 11th base, leaving the entire reads. Quality control and preprocessing of FASTQ files were performed using fastp version 0.20.1 ^12^. Each FASTQ per sample was mapped to SARS-CoV-2 reference genome (GenBank no. MN908947) using bwa version 0.7.17-r1188 ^13^. BAM files were sorted by SAMtools version 1.9 ^14^, and variants were called using bcftools version 1.9 ^15^, filtered by quality-measure of at least 20 and read depth of at least 50.

#### Machine learning models

Classification models were implemented using scikit-learn module in Python, version 0.23.2 ^16^. For the initial model, the samples were randomly divided into 60% train set, 20% validation set and 20% test set. SARS-COV-2 genes counts, normalized for total read number per sample, were selected as features, while the label of a positive/negative diagnosis was based on traditional RT-PCR. Hyperparameters were selected after an exhaustive grid search, with 5-fold cross validation. Selected hyperparameters were those which provided the maximal Area Under the Receiver Operating Characteristic (ROC) curve (AUC) scores. Then, the model was evaluated by predicting the diagnosis in the test set. Additional models, i.e., based on different gene combinations, were methodologically similar.

## Data Availability

Additional data can be available upon request.

